# Life-course burden of health deficits associates with later-life heart size and function in the 1946 British birth cohort

**DOI:** 10.1101/2020.08.13.20174599

**Authors:** Constantin-Cristian Topriceanu, James C Moon, Rebecca Hardy, Nish Chaturvedi, Alun D Hughes, Gabriella Captur

**Affiliations:** UCL Medical School, 74 Huntley St, Bloomsbury, London WC1E 6DE; UCL MRC Unit for Lifelong Health and Ageing, University College London, Fitzrovia, London WC1E 7HB; UCL Institute of Cardiovascular Science, University College London, Gower Street, London WC1E 6BT, UK; Cardiac MRI Unit, Barts Heart Centre, West Smithfield, London, EC1A 7BE; CLOSER, UCL Institute of Education, 55-59 Gordon Square, London WC1H 0NU; The Royal Free Hospital, Centre for Inherited Heart Muscle Conditions, Cardiology Department, Pond Street, Hampstead, London NW3 2QG, UK

**Keywords:** frailty, cardiovascular disease, ejection fraction, left ventricular mass index, myocardial contraction fraction, E/e’

## Abstract

**Aim:** To study the association between the life-course accumulation of health deficits and later-life heart size and function using data from the 1946 National Survey of Heath and Development (NSHD) British birth cohort, the world’s longest running birth cohort with continuous follow-up.

**Methods and Results:** A multidimensional health deficit index (DI) looking at 45-health deficits was serially calculated at 4 time periods of the life-course in NSHD participants (0-16, 19-44, 45-54 and 60 to 64 years), and from these the mean and total DI for the life-course was derived (DI_mean_, DI_sum_). The step change in deficit accumulation from one time period to another was also calculated. Echocardiographic data at 60-64 years provided: ejection fraction (EF), left ventricular mass indexed to body surface area (LVmassi, BSA), myocardial contraction fraction indexed to BSA (MCF_i_) and E/e’. Generalized linear models assessed the association between DIs and echocardiographic parameters after adjustment for sex, socio-economic position and body mass index.

1,375 NSHD participants were included (46.47% male). For each single new deficit accumulated at any one of the 4 time periods of the life-course, LVmass_i_ increased by 0.91-1.44% (*p*<0.013), while MCF_i_ decreased by 0.6-1.02% (*p*<0.05 except at 45-54 years). One unit increase in DI at age 45-54 and 60-64 decreased LV EF by 11-12% (*p*<0.013). A single deficit step change occurring between 60-64 years and one of the earlier time periods, translated into significantly higher odds (2.1—78.5, *p*<0.020) of elevated LV filling pressure defined as E/e’>13.

**Conclusion:** The accumulation of health deficits at any time period of the life-course associates with a maladaptive cardiac phenotype in older age, dominated by myocardial hypertrophy and poorer function. The burden of health deficits appears to strain the myocardium potentially leading to future cardiac dysfunction.

## INTRODUCTION

Aging is an inevitable process of biological life. In general, as humans age they accumulate more health deficits which are translated into adverse health outcomes, complex medical needs and high financial burden on society^1^. However, people do not age and accumulate health deficits at the same rate, highlighting the difference between chronological and biological age.

The accumulation of new health deficits throughout life results in a multidimensional loss of reserves spanning the physical and cognitive domains. This health deficit burden in an individual can be quantified and there are mainly three schools of thought for how to do this: 1) the rules-based operational definition approach relies on a set of phenotypical rules of performance against which the individual is scored^2^; 2) the clinical frailty score grades persons along a spectrum from very fit to very frail based on clinical judgment; and 3) the health deficit index (DI, also known as frailty index) counts health deficit accumulation and it is the ratio between the number of deficits in an individual, and the total number of deficits appraised, with sub-unitary scores for partial deficits^3^. The DI is most widely used in research because it is easy to score, it has the mathematical advantage of accommodating partial deficits, and it correlates with the clinical frailty score^4^. The DI should not be confused with a physical frailty phenotypic score that only appraises domains such as shrinking, exhaustion, low physical activity, slow gait speed and weak grip strength. The DI has been extensively validated and shown to predict mortality^5^ and other negative health outcomes^6^.

Cardiovascular diseases (CVDs) are an important component of the multi-morbidity syndrome accounting for more than 30% of all global deaths. Besides the traditional risk factors such as diabetes and hypertension, it is not known whether multi-morbidity and its quantifiable equivalent, the DI, are related to the appearance of CVD. It is also unknown whether a greater burden of health deficits in early-life predicts later-life CVD. To answer these questions, we analyzed life-course data of participants from the 1946 Medical Research Council (MRC) British National Survey of Health and Development (NSHD)-the world’s longest running birth cohort with continuous follow-up. We calculated DIs at different periods of the life-course and examined their association with cardiac size and function at age 60-64 years. The importance of the 7^th^ decade stems from the current 80-year human life-expectancy-having cardiac dysfunction at 60 translates into either premature death or living two further decades but with a lower quality of life.

## METHODS

### Study design

Participants were from the MRC NSHD, a birth cohort study comprised of 5,362 individuals born in 1 week in 1946 in Britain. The cohort has been extensively followed up with periodic assessments which have been described elswehere^7^. The 2006-2010 NSHD data collection was granted ethical approval from the Greater Manchester Local Research Ethics Committee and the Scotland Research Ethics Committee. Participants’ weight and height were measured at 60-64 years and used to compute body mass index (BMI) and body surface area (BSA). Participants’ socioeconomic position (SEP) was evaluated at the time of echocardiography (60-64 years) or at 53 years where the former was not available, according to the UK Office of Population Censuses and Surveys Registrar General’s social class.

### Construction of the health deficit index

The definition of a health deficit is broad and includes symptoms, signs, medical conditions, disabilities, laboratory and imaging abnormalities^8^. In line with the Rockwood approach^3^ deficits included in the DI (**Figure 1, Supplementary Tables S1-S3**) had to meet the following criteria: health related; prevalence between 1% to 80%; span a wide variety of medical systems; contain less than 5% missing data^4^. To sequentially apply the same index on cohort participants at different time points, we appraised the same health deficits at each period of the life-course. For reliability, we aimed for an index with at least 30 health deficits in line with previous works^3^.

**Figure 1.**
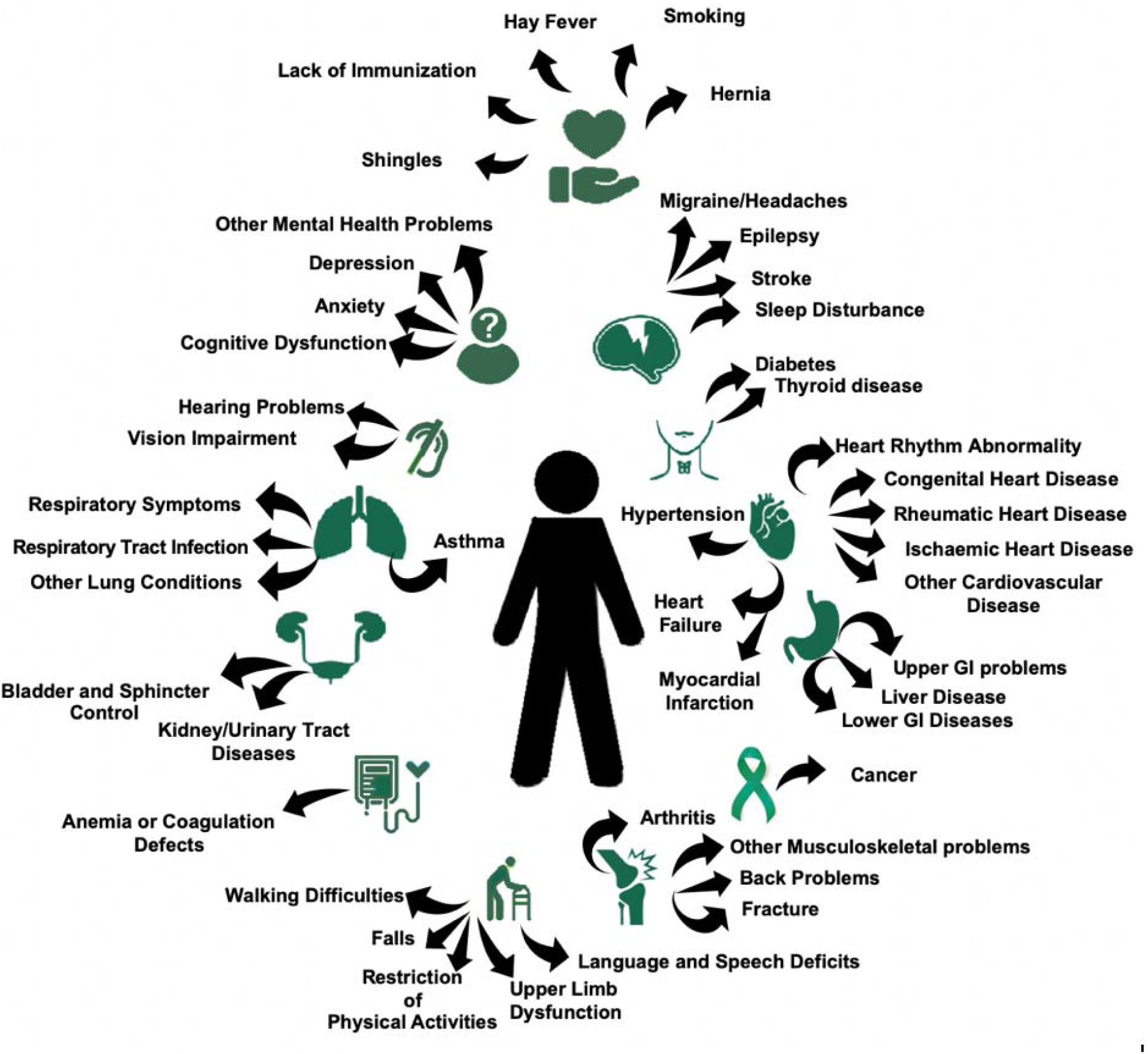
The 45 health deficit categories, spanning a broad set of physical and cognitive domains, appraised in MRC NSHD 1946 British birth cohort participants. *MRC NSHD, Medical Research Council National Survey of Health and Development; GI, gastrointestinal*.

Four time periods were examined: 0 to 16 years representing early-life (DI_0_16_), 19 to 44 years representing young adulthood (DI _19_44_), 45 to 54 representing early rmiddle age (DI_45_54_) and 60 to 64 representing older age (DI_60_64_).

Health deficit score was “0” for complete absence, “1” for absolute presence, and “0.5” for partial deficits. Irreversible medical events (e.g. stroke) carried their deficit score onto all subsequent time periods, while reversible events (e.g. bone fracture) were scored *de novo* at each time point. To calculate the DI, the sum of health deficits present in an individual was divided by the total number of possible deficits in our index (that is 45) minus the number of missing deficits for that participant (**Equation 1**). The resultant DI can be a number from 0 (least) to 1 (most).

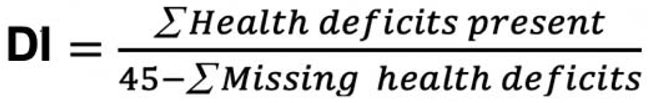

Equation 1: Formula for calculating the DI.

To assess whole-of-life health deficit burden the 4 individual DIs were averaged (DI_mean_) and summed (DI_sum_). To assess health deficit change between time periods, the difference between DIs at the 4 time points were computed as follows: DI_2–1_=(DI_19___44_)-(DI_0_16_); DI_3_1_ = (DI_45_54_)-(DI_0_16_); DI_4_1_ = (DI_60_64_)-(DI_0_16_); DI_3_2_ = (DI_45_54_)-(DI_19_44_); D_14_2_ = (DI_60_64_)-(DI_19_44_); and DI_4–3_=(DI_60_64_)-(DI_45_54_).

### Echocardiography

Between 2006-2010 when study members were 60-64 years, British-based NSHD participants who had not been lost to follow-up or withdrawn, were invited to attend a clinic-based assessment that included resting transthoracic echocardiography using General Electric (GE) Vivid I machines. Briefly, the echocardiographic protocol^7^ included long and short axis parasternal views, apical 5-, 4, 3- and 2-chamber views, aortic short axis view, and conventional/tissue Doppler in the apical 4-chamber. Reproducibility between readers was previously reported (intraclass correlation coefficients >0.80)^9^. Ejection Fraction (EF) was calculated by the biplane Simpson’s method. Left ventricular (LV) mass was indexed by BSA to obtain the LV mass index (LVmass_i_) as per the American Society of Echocardiography recommendations. Myocardial contraction fraction index (MCF_i_) was calculated as the ratio between stroke volume and myocardial volume. Myocardial volume was calculated by dividing LVmass_i_ by mean myocardial density (1.04 g/ml)^10^. E/e’ ratio was calculated by dividing peak mitral valve velocity (E) by the average of lateral and septal mitral annular early diastolic velocity (e’)^11^. LV end-diastolic volume index (LVEDV_I_) was derived by dividing LVEDV by BSA.

### Statistics

Statistical analysis was performed in R (version-3.6.0). Distribution of data were assessed on histograms and using Shapiro-Wilk test. Continuous variables are expressed as mean ± 1 standard deviation (SD); categorical variables, as counts and percent. Regression models were developed using DIs or their derivatives as exposures to predict LV EF, LVmass_i_, MCF_i_ and E/e’ by echocardiography at 60-64 years. Model 1 was adjusted for sex, Model 2 for sex and SEP, and Model 3 for sex, SEP and BMI. For LVmassi and MCFi, anthropometry was already accounted for through BSA indexation, so BMI adjustment was not additionally pursued. This decision during study design was informed through the use of directed acrylic graphs (visual representations of causal assumptions that can identify sources of confounding) that helped identify potential colliders (in this case BMI) that should be left uncontrolled, versus non-colliders (or confounders) that should be controlled in the regression models^12^.

As a result of the skewed distribution of echocardiographic parameters, generalized linear models (glm) with gamma distribution and log link were used to investigate the association of DIs with the following continuous outcome variables: LV EF, LVmass_i_ and MCF_i_; while glm with binomial distribution and logit link (equivalent to logistic regression) was employed for the binary outcome variable E/e’ using a cut-off of >13. A two-tailed *p*-value <0.05 was considered statistically significant. Model assumptions were verified with regression diagnostics to ascertain linearity, normality, homoscedasticity and independence of the residuals.

We ran sensitivity analyses in which we: (1) assessed whether the associations of DI scores with echocardiographic parameters remained unchanged if all cardiovascular-related health deficits were excluded from the DIs (we removed hypertension, ischaemic heart disease, myocardial infarction, heart failure, heart rhythm abnormality, congenital heart disease, rheumatic heart disease, and other cardiovascular diseases); (2) assessed whether additional adjustment for childhood SEP (over and above adjustment for sex, BMI and late-adulthood SEP) was pursued; (3) re-analyzed associations between DIs and EF/LVmass_i_ as binary rather than continuous outcome variables using an EF cut-off of 55% and LVmass_i_ cut-offs of 102 and 88g/m^2^ for men and women respectively, as per British Society of Echocardiography Guidelines for Chamber Quantification^13^; (4) compared the echocardiographic parameters at age 60-64 of excluded participants with >20% missing data (for which DIs were not calculated) vs. included participants who had <20% of missing data; (5) compared participants with complete echocardiographic variables and those with one or more missing echocardiographic parameter; (6) simulated a complete case analysis in terms of the outcome variables, through multiple imputation. To verify the reliability of the health deficit index, we performed a principal component analysis (PCA) of the included health deficits. We constructed a PCA-weighted health deficit index (wDI) via the weighted average of the health deficits using the first principal component weights (**Supplementary Table S2**). This procedure was done for two age-interval exemplars: young adulthood (19 to 44 years) and older age (60 to 64 years). For each of these wDIs, we then re-ran the fully adjusted regression models using wDI_19___44_, and wDI_60___64_ respectively. To check for different profiles in the scores, we performed a multiple correspondence analysis (MCA) based on the indicator matrix. A scree plot was used to extract the optimal number of dimensions and to determine the percentages of inertia. From the geometric space created in the MCA, participants were then classified into clusters according to proximity criteria using the k-means algorithm. This separated participants into homogenous groups while maximizing heterogeneity across groups.

## RESULTS

### Participant characteristics and life-course burden of health deficits

Ninety health deficits were initially considered (**Supplementary Table S1**). Forty-five health deficits met the criteria for inclusion in the DI (**Figure 1, Supplementary Table S2-S3**). Of the 2,856 participants invited to attend the clinic visit at 60-64 years, 1,690 attended and 1,653 had echocardiography of which 1,617 had acceptable image quality. Of these, 1,375 study members had <20% missing data and at least one outcome parameter of interest by echocardiography. Out of the 1375 NSHD participants studied, 1 had an DI=0 plus missing health deficit data of up to 20%, while 3 had DI=0 in the absence of missing data.

Participant characteristics and DI results are summarized in **Table 1**. Generally, higher DI_sum_ and DI_mean_ were associated with female sex, lower SEP and higher BMI.

**Table 1.** Characteristics of the MRC NSHD sample considering participants with less than <20% missing health deficit data and having at least one echocardiographic parameter of interest (EF, MCF_i_, LVmass_i_ or E/e’).

|  | Overall |  | Men |  | Women |  |
| --- | --- | --- | --- | --- | --- | --- |
|  | *n* | Result | *n* | Result | *n* | Result |
| Age, years | 1375 | 63.2 ± 1.1 | 639 | 63.2 ± 1.2 | 736 | 63.3 ± 1.1 |
| Height, m | 1375 | 1.68 ± 0.88 | 639 | 1.75 ± 0.06 | 736 | 1.62 ± 0.06 |
| Weight, kg | 1375 | 78.13 ± 14.80 | 639 | 84.60 ± 12.79 | 736 | 73.16 ± 14.66 |
| BMI | 1375 | 27.62 ± 4.63 | 639 | 27.63 ± 3.91 | 736 | 28.03 ± 5.59 |
| SEP Non-manual * | 1375 | 1003 (72.94) | 639 | 436 (63.31) | 736 | 567 (77.04) |
| SEP Manual<sup>§</sup> | 1375 | 372 (31.76) | 639 | 203 (31.77) | 736 | 169 (22.96) |
| LVEDV<sub>i</sub> | 1222 | 48.25 ± 11.18 | 579 | 52.18 ± 12.6 | 643 | 44.72 ± 8.27 |
| IVSD, cm | 1256 | 1.06 ± 0.25 | 581 | 1.12 ± 0.24 | 675 | 1.01 ± 0.24 |
| PWT, cm | 1254 | 0.99 ± 0.21 | 580 | 1.06 ± 0.24 | 674 | 0.94 ± 0.17 |
| RWT | 1254 | 0.44 ± 0.11 | 580 | 0.44 ± 0.11 | 674 | 0.43 ± 0.11 |
| LAVI, ml/m<sup>2</sup> | 1195 | 22.08 ± 7.59 | 571 | 23.13 ± 7.90 | 624 | 21.12 ± 7.16 |
| EF, % | 1217 | 64.41 ± 7.89 | 576 | 63.45 ± 7.07 | 640 | 65.28 ± 7.63 |
| LVmass<sub>i</sub>, g/m<sup>2</sup> | 1006 | 114.91 ± 37.50 | 448 | 129.38 ± 41.30 | 558 | 103.30 ± 29.39 |
| MCF<sub>i</sub> | 860 | 0.49 ± 0.19 | 395 | 0.52 ± 0.20 | 465 | 0.48 ± 0.18 |
| E/e' | 1259 | 7.95 ± 2.13 | 569 | 7.51 ± 2.03 | 690 | 8.3 ± 2.15 |
| DI<sub>0_16</sub> | 1375 | 0.14 ± 0.05 | 639 | 0.13 ± 0.05 | 736 | 0.14 ± 0.05 |
| DI<sub>19_44</sub> | 1375 | 0.14 ± 0.07 | 639 | 0.13 ± 0.07 | 736 | 0.15 ± 0.07 |
| DI<sub>45_54</sub> | 1375 | 0.21 ± 0.08 | 639 | 0.19 ± 0.07 | 736 | 0.24 ± 0.08 |
| DI<sub>60_64</sub> | 1375 | 0.28 ± 0.09 | 639 | 0.25 ± 0.09 | 736 | 0.29 ± 0.09 |
| DI<sub>2-1</sub> | 1375 | 0.003 ± 0.07 | 639 | -0.005 ± 0.06 | 736 | 0.009 ± 0.07 |
| DI<sub>3-1</sub> | 1375 | 0.08 ± 0.08 | 639 | 0.05 ± 0.07 | 736 | 0.10 ± 0.08 |
| DI<sub>4-1</sub> | 1375 | 0.14 ± 0.09 | 639 | 0.12 ± 0.09 | 736 | 0.16 ± 0.09 |
| DI<sub>3-2</sub> | 1375 | 0.07 ± 0.06 | 639 | 0.05 ± 0.05 | 736 | 0.09 ± 0.06 |
| DI<sub>4-2</sub> | 1375 | 0.14 ± 0.07 | 639 | 0.1 ± 0.07 | 736 | 0.15 ± 0.08 |
| DI<sub>4-3</sub> | 1375 | 0.06 ± 0.06 | 639 | 0.07 ± 0.06 | 736 | 0.05 ± 0.06 |
| DI<sub>sum</sub> | 1375 | 0.77 ± 0.23 | 639 | 0.70 ± 0.21 | 736 | 0.82 ± 0.24 |
| DI<sub>mean</sub> | 1375 | 0.19 ± 0.06 | 639 | 0.18 ± 0.05 | 736 | 0.21 ± 0.06 |

\*Defined as SEP classes I-IIIIN.
<sup>§</sup>Defined as SEP classes IIIM-V.
Data presented as mean ± standard deviation or counts (%) as appropriate.

For the vast majority of participants DI increased across the life-course, but in 86 participants (0.05%) DI decreased in spite of aging, largely due to the reversible nature of some health deficits. There was no significant difference in echocardiographic parameters between participants whose life-course DIs decreased when compared to the rest of the cohort (LV EF *p*=0.30, LVmass_i_ *p*=0.99, MCF_i_ *p*=0.08, E/e’ *p*=0.57).

There was no difference in Di between the ages 0-16 and 19-44 years, but increases were observed between young adulthood and early middle age, and between early middle age and older age. Raw health deficit index scores of NSHD participants are displayed in **Supplementary Figure S1** and the life-course burden of health deficits in **Figure 2**. Health deficit contributors at the 4 time periods are shown in **Supplementary Figure S2**. Women consistently had higher DIs than men at every age interval (**Figure 3**).

**Figure 2.**
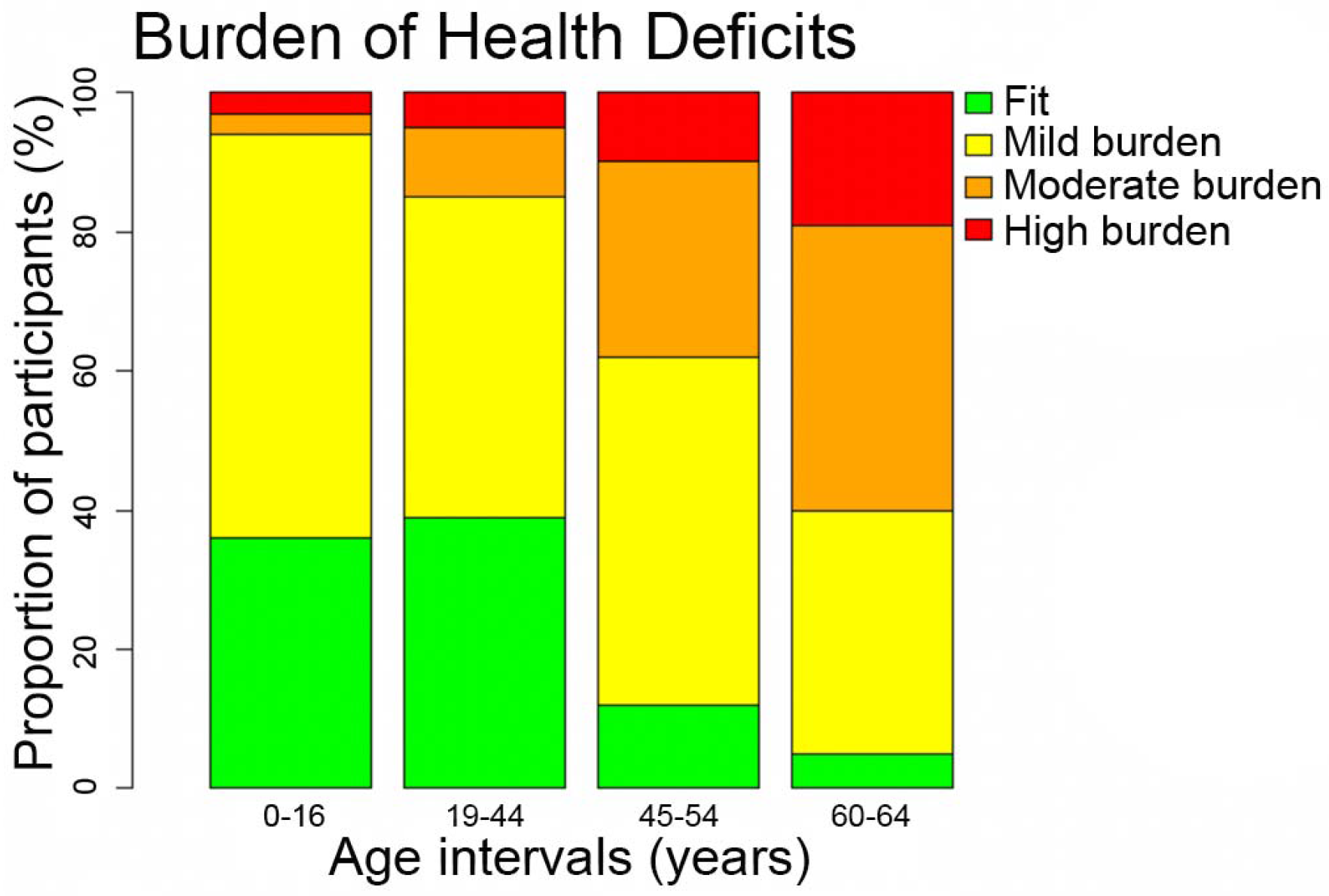
Bar chart illustrating the life-course burden of health deficits in NSHD participants. Health deficit categories have been defined as follows: fit, DI<0.12; low burden, 0.12≤DI<0.24; moderate burden, 0.24≤DI<0.36; high burden, 0.36≤DI. *DI, health deficit index*.

**Figure 3.**
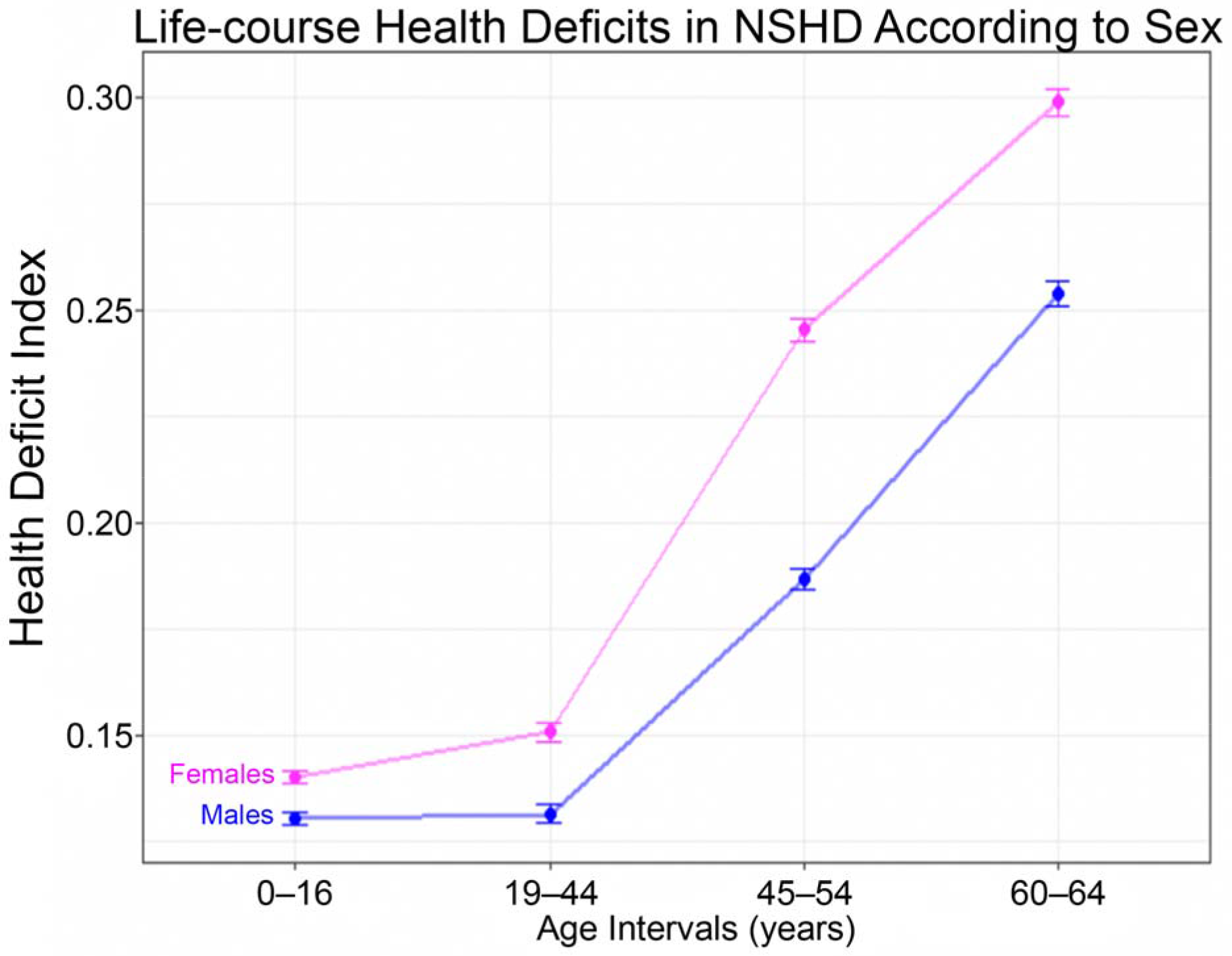
Sex differences in life-course burden of health deficits in the NSHD cohort. Females (pink) had higher DIs (y-axis) than males (blue) at every time period (x-axis). Vertical bars represent standard error of the means. Abbreviations as in Figure 2.

### Ejection fraction

The association between DIs and echocardiographic parameters are presented in **Table 2**. In fully adjusted models, a unit increase in DI (from 0 to 1) in early middle age (DI45_54) or older age (DI60_64) translated into an 11-12% decrease in EF (p=0.013 and 0.002). A unit increase in DIsum and DImean translated into a 4-15% decrease in EF (p=0.011 both). When comparing the DIs of participants with the highest and lowest deciles of EF, those with the lowest EF had higher DIs throughout life except in early life (0-16 years) (**Figure 4A**).

**Table 2.** Associations between health deficit burden at the 4 time periods, step-change in deficit accumulation and whole-of-life health deficit burden, with echocardiographic parameters (EF, LVmass_i_, MCF_i_ and E/e’) at 60-64 years.

|  |  | **Model 1**<br>**(adjusted for sex)** |  | **Model 2**<br>**(adjusted for sex + SEP)** |  | **Model 3**<br>**(adjusted for sex + SEP + BMI)** |  |
| --- | --- | --- | --- | --- | --- | --- | --- |
| **Echo Parameter** | **DI** | **$\beta$ (95% CI)** | ***p*-value** | **$\beta$ (95% CI)** | ***p*-value** | **$\beta$ (95% CI)** | ***p*-value** |
| **EF\*** | DI<sub>0_16</sub> | 0.03 (-0.12, 0.18) | 0.667 | 0.03 (-0.12, 0.17) | 0.745 | 0.04 (-0.11, 0.19) | 0.623 |
|  | DI<sub>19_44</sub> | -0.10 (-0.20, 0.00) | 0.058 | -0.11 (-0.21, 0.00) | 0.055 | -0.10 (0.20, 0.01) | 0.070 |
|  | DI<sub>45_54</sub> | -0.12 (-0.21, -0.03) | **0.010** | -0.12 (-0.21, -0.03) | **0.007** | -0.12 (-0.21, -0.03) | **0.013** |
|  | DI<sub>60_64</sub> | -0.12 (-0.20, -0.05) | **0.001** | -0.13 (-0.21, -0.06) | **<0.001** | -0.13 (-0.20, -0.05) | **0.002** |
|  | DI<sub>2-1</sub> | -0.12 (-0.22, -0.01) | **0.028** | -0.12 (-0.22, -0.01) | **0.025** | -0.11 (-0.22, -0.01) | **0.032** |
|  | DI<sub>3-1</sub> | -0.13 (-0.21, -0.04) | **0.005** | -0.13 (-0.22, -0.04) | **0.004** | -0.12 (-0.21, -0.03) | **0.007** |
|  | DI<sub>4-1</sub> | -0.13 (-0.21, -0.06) | **<0.001** | -0.14 (-0.21, -0.06) | **<0.001** | -0.13 (-0.21, -0.06) | **0.001** |
|  | DI<sub>3-2</sub> | -0.08 (-0.20, 0.04) | 0.210 | -0.08 (-0.20, 0.04) | 0.194 | -0.07 (-0.19, 0.05) | 0.224 |
|  | DI<sub>4-2</sub> | -0.11 (-0.20, -0.01) | **0.025** | -0.11 (-0.21, -0.02) | **0.019** | -0.11 (-0.20, -0.01) | **0.027** |
|  | DI<sub>4-3</sub> | -0.10 (-0.21, 0.02) | 0.115 | -0.10 (-0.22, 0.02) | 0.099 | -0.10 (-0.21, 0.02) | 0.117 |
|  | DI<sub>sum</sub> | -0.04 (-0.07, -0.01) | **0.009** | -0.04 (-0.07, -0.01) | **0.006** | -0.04 (-0.07, -0.01) | **0.011** |
|  | DI<sub>mean</sub> | -0.16 (-0.28, -0.04) | **0.009** | -0.17 (-0.29, -0.05) | **0.006** | -0.16 (-0.28, -0.04) | **0.011** |
| **LVmass<sub>i</sub>** | DI<sub>0_16</sub> | 0.58 (0.18, 0.97) | **0.005** | 0.50 (0.11, 0.90) | **0.013** |  |  |
|  | DI<sub>19_44</sub> | 0.46 (0.19, 0.74) | **0.001** | 0.41 (0.14, 0.69) | **0.003** |  |  |
|  | DI<sub>45_54</sub> | 0.40 (0.16, 0.63) | **<0.001** | 0.34 (0.11, 0.58) | **0.004** |  |  |
|  | DI<sub>60_64</sub> | 0.40 (0.20, 0.61) | **<0.0001** | 0.36 (0.16, 0.56) | **<0.001** |  |  |
|  | DI<sub>2-1</sub> | 0.19 (-0.09, 0.47) | 0.183 | 0.17 (-0.11, 0.45) | 0.221 |  |  |
|  | DI<sub>3-1</sub> | 0.20 (-0.04, 0.45) | 0.094 | 0.17 (-0.07, 0.41) | 0.157 |  |  |
|  | DI<sub>4-1</sub> | 0.26 (0.06, 0.47) | **0.012** | 0.23 (0.03, 0.44) | **0.025** |  |  |
|  | DI<sub>3-2</sub> | 0.12 (-0.21, 0.45) | 0.467 | 0.08 (-0.24, 0.41) | 0.618 |  |  |
|  | DI<sub>4-2</sub> | 0.25 (-0.01, 0.50) | 0.063 | 0.21 (-0.04, 0.47) | 0.102 |  |  |
|  | DI<sub>4-3</sub> | 0.27 (-0.05, 0.59) | 0.102 | 0.26 (-0.06, 0.58) | 0.115 |  |  |
|  | DI<sub>sum</sub> | 0.17 (0.09, 0.25) | **<0.0001** | 0.15 (0.07, 0.23) | **<0.001** |  |  |
|  | DI<sub>mean</sub> | 0.67 (0.36, 0.99) | **<0.0001** | 0.60 (0.28, 0.91) | **<0.001** |  |  |
| **MCF<sub>i</sub>** | DI<sub>0_16</sub> | -0.66 (-1.23, 0.09) | **0.023** | -0.61 (-1.18, -0.03) | **0.039** |  |  |
|  | DI<sub>19_44</sub> | -0.53 (-0.91, -0.14) | **0.008** | -0.48 (-0.87, -0.09) | **0.016** |  |  |
|  | DI<sub>45_54</sub> | -0.38 (-0.69, -0.06) | **0.025** | -0.32 (-0.65, -0.001) | 0.054 |  |  |
|  | DI<sub>60_64</sub> | -0.43 (-0.70, -0.15) | **0.003** | -0.38 (-0.66, -0.10) | **0.009** |  |  |
|  | DI<sub>2-1</sub> | -0.23 (-0.62, 0.16) | 0.256 | -0.20 (-0.60, 0.19) | 0.310 |  |  |
|  | DI<sub>3-1</sub> | -0.17 (-0.50, 0.16) | 0.312 | -0.13 (-0.46, 0.20) | 0.431 |  |  |
|  | DI<sub>4-1</sub> | -0.27 (-0.55, 0.01) | 0.064 | -0.23 (-0.52, 0.06) | 0.113 |  |  |
|  | DI<sub>3-2</sub> | -0.02 (-0.48, 0.43) | 0.922 | 0.02 (-0.44, 0.48) | 0.938 |  |  |
|  | DI<sub>4-2</sub> | -0.24 (-0.60, 0.12) | 0.182 | -0.20 (-0.56, 0.17) | 0.276 |  |  |
|  | DI<sub>4-3</sub> | -0.36 (-0.82, 0.10) | 0.116 | -0.33 (-0.79, 0.13) | 0.151 |  |  |
|  | DI<sub>sum</sub> | -0.18 (-0.29, -0.07) | **0.002** | -0.16 (-0.27, -0.05) | **0.006** |  |  |
|  | DI<sub>mean</sub> | -0.71 (-1.14, -0.27) | **0.002** | -0.63 (-1.07, -0.19) | **0.006** |  |  |
| **E/e'** | DI<sub>0_16</sub> | 4.70 (-3.27, 12.39) | 0.239 | 4.76 (-3.22, 12.45) | 0.234 | 2.72 (-5.31, 10.72) | 0.489 |
|  | DI<sub>19_44</sub> | 0.77 (-5.01, 6.22) | 0.788 | 0.81 (-4.99, 6.28) | 0.778 | -0.82 (-6.83, 4.84) | 0.782 |
|  | DI<sub>45_54</sub> | 4.28 (-0.35, 8.65) | 0.062 | 4.39 (-0.28, 8.82) | 0.058 | 2.87 (-1.99, 7.49) | 0.235 |
|  | DI<sub>60_64</sub> | 6.39 (2.62, 10.06) | **<0.001** | 6.50 (2.71, 10.18) | **<0.001** | 5.26 (1.37, 9.06) | **0.007** |
|  | DI<sub>2-1</sub> | -1.61 (-7.45, 3.99) | 0.581 | -1.60 (-7.44, 4.00) | 0.584 | -2.25 (-8.23, 3.46) | 0.449 |
|  | DI<sub>3-1</sub> | 2.71 (-2.04, 7.22) | 0.250 | 2.75 (-2.01, 7.28) | 0.245 | 1.84 (-3.04, 6.49) | 0.449 |
|  | DI<sub>4-1</sub> | 5.39 (1.53, 9.13) | **0.006** | 5.44 (1.57, 9.17) | **0.005** | 4.55 (0.64, 8.33) | **0.020** |
|  | DI<sub>3-2</sub> | 7.08 (0.71, 13.30) | **0.027** | 7.15 (0.75, 13.42) | **0.027** | 6.31 (-0.25, 12.74) | 0.056 |
|  | DI<sub>4-2</sub> | 9.31 (4.64, 13.90) | **<0.0001** | 9.36 (4.69, 13.96) | **<0.0001** | 8.13 (3.48, 12.78) | **<0.001** |
|  | DI<sub>4-3</sub> | 9.00 (2.74, 15.16) | **0.004** | 9.09 (2.81, 15.29) | **0.004** | 8.17 (1.99, 14.33) | **0.009** |
|  | DI<sub>sum</sub> | 1.78 (0.23, 3.27) | **0.021** | 1.82 (0.26, 3.33) | **0.019** | 1.26 (-0.36, 2.83) | 0.119 |
|  | DI<sub>mean</sub> | 7.12 (0.91, 13.07) | **0.021** | 7.30 (1.04, 13.31) | **0.019** | 5.05 (-1.45, 11.31) | 0.119 |

\* All reported analyses here consisted of generalized linear models with gamma distribution and log link except for E/e' that used logistic regression.
Significant *p*-values are highlighted in bold.
β, regression coefficient; CI, confidence interval. Other abbreviations as in **Table 1**.

**Figure 4.**
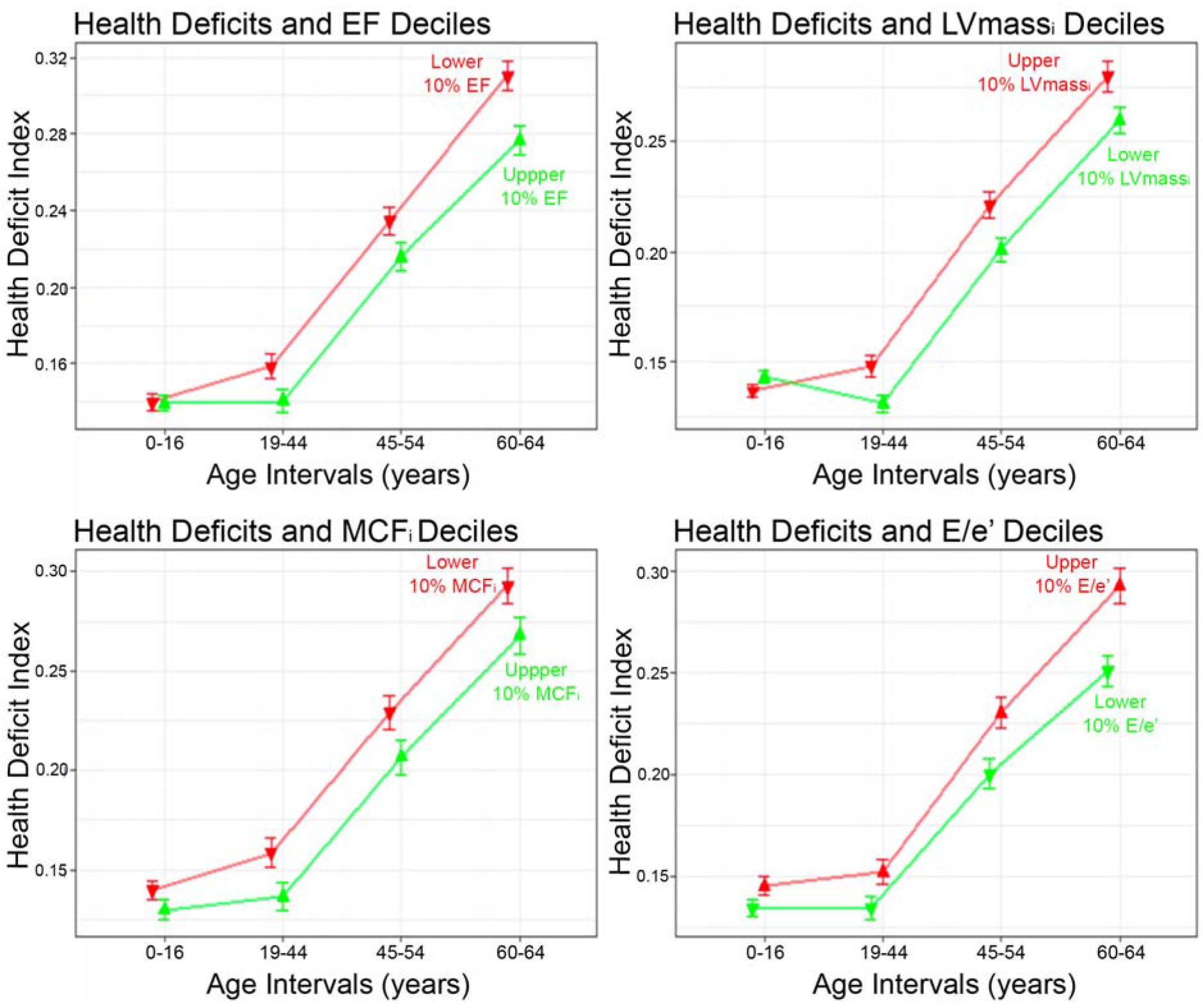
Comparing trajectories of mean life-course DI (*y*-axis) at each age interval (*x*-axis) in terms of the older age echocardiographic phenotype. DI trajectories leading to favourable phenotypes shown in green and unfavourable trajectories in red as follows: (**A**) highest (green) vs. lowest (red) LV EF deciles; (**B**) highest (red) vs. lowest (green) LVmass_i_ deciles; (**C**) highest (green) vs. lowest (red) MCF_i_ deciles; and (**D**) highest (red) vs. lowest (green) E/e’ deciles. Vertical bars represent standard error of the means. Wilcoxon signed-rank test p-values (not shown) for median inter-decile differences were <0.05 at every time point except for the 0-16 years age interval. *EF, ejection fraction; LVmassi, left ventricular mass index; MCFi, myocardial contraction fraction index*. Other abbreviations as in **Figure 2**.

### Left ventricular mass index

In fully adjusted models, a unit increase in DIs translated into a 16-82% increase in LVmass_i_ (all p<0.013). The accumulation of a single new health deficit in early-life, young adulthood, early middle age or older age, led to a 1.44%, 1.13%, 0.91% and 0.96% increase in LVmassi respectively. The group of participants with the highest decile of LVmass_i_ had significantly higher DIs throughout life when compared to the group with the lowest decile, except in early-life (**Figure 4B**).

### Myocardial contraction fraction index

In fully adjusted models, a unit increase in DI translated into a 15%-47% decrease in MCF_i_ (all *p*<0.05). The accumulation of a single new health deficit in early-life, young adulthood, early middle age or older age, led to a 1.02%, 0.85%, 0.60% and 0.71% decrease in MCF_i_ respectively. When comparing the DIs of participants with the highest and lowest deciles of MCF_i_, those with the lowest MCF_i_ had significantly higher DIs throughout life except in early-life (**Figure 4C**).

### E/e’ ratio

A unit increase in DI in older age increased the log odds of having elevated LV filling pressure (E/e’ >13) by 5.3 (*p*=0.007) in fully adjusted models. The addition of one new health deficit from early-life to older age (DI_4–1_) increased the multiplicative odds of having elevated LV filling pressure by 2.1, by 75.4 times for DI_4–2_ and 78.5 times for DI_4–3_. Participants with the highest decile of E/e’ had significantly higher DIs throughout life when compared to those in the lowest decile (**Figure 4D**).

### LVEDV_i_

After adjustment for sex and SEP there was no association between any of the DIs and LVEDV_i_ (**Supplementary Table S4**).

### Sensitivity analysis

After removing cardiovascular health deficits from all the DIs, associations of early-life and/or young adulthood health deficit burden with the 4 key echocardiographic parameters, remained significant for LVmass_i_ and MCF_i_ (**Supplementary Table S5**). After additional adjustment for childhood SEP, *β* coefficients were slightly attenuated but DIs mostly retained significant associations with echocardiographic parameters (**Supplementary Table S6**). There was no significant difference (*p*>0.10) between the echocardiographic parameters of participants included in the DI analysis compared to those excluded due to >20% missing data (**Supplementary Table S7**). There was no significant difference in cardiac values (*p*>0.50) between those with complete or some missing echocardiographic parameters (**Supplementary Table S8**). Associations between DIs and echocardiographic parameters persisted in fully adjusted models after multiple imputation (**Supplementary Table S9**). Associations between PCA *w*DIs and LVmass_i_, and MCF_i_ persisted both in young adulthood and older age, but were lost for EF and E/e’ (**Supplementary Table S10**). The cluster analysis revealed no clear separation of data in terms of health deficit patterns in any of the age-intervals. (**Supplementary Figures S4-S7**).

## DISCUSSION

Life-course data from the MRC NSHD British birth cohort indicates that health deficits accumulated during childhood, young adulthood, middle age and older age, and the accumulation of new health deficits between these periods, associate with an adverse cardiac phenotype in older age–one that is characterized by hypertrophy, elevated filling pressure and reduced systolic function.

A health deficit index objectively captures the vulnerability of an individual by counting health deficits. Although theoretically a DI can have any value from 0 to 1, values higher than 0.5 were rare amongst NSHD participants (<1%), as a high burden of health deficits limits survival^14^. The relatively large proportion of individuals at 0-16 and 19-44 years with low health deficit burden (**Figure 2**) can be explained by the broad time periods over which health deficits were scored (~20 years) as well as the existence of broadly defined health deficits encompassing multiple clinical entities (**Supplementary Table S3**).

The burden of health deficits in all 4 time periods of the life-course, including early-life and young adulthood, was associated with higher LVmass_i_ and lower MCF_i_ in later-life (60-64 years). A possible explanation may be that health deficits insidiously strain the heart which eventually manifests as maladaptive myocardial hypertrophy. Even after removing all cardiovascular-related health deficits from the DIs, the associations remained significant, particularly in early-life, confirming the biological relationship linking any kind of global index of health deficits with cardiac function parameters in older age.

We found that NSHD participants with decreasing DIs across the life-course, had fitter hearts (higher EF) when compared to the rest of the cohort. Recognizing the potential long-term cardiovascular sequelae of health deficit accrual in these early formative years opens a window of opportunity for managing health-deficits earlier rather than later. This means firmer implementation of public health strategies aimed at disease prevention, the integrated treatment of conditions as soon as they arise, and provision of more holistic and integrated models of healthcare to ensure that optimal care is provided for all co-existing conditions. Importantly, there is a need to ensure that the priorities and preferences of patients are integrated in all of these strategies to maximize what may well become their life-course engagement with the healthcare system.

Whole-of-life burden of health deficits (DI_sum_ and DI_mean_) was associated with lower EF and MCF_i_, and higher LVmass_i_ in later-life, more so for DI_mean_. The latter is likely due to the over-inflation of the DI brought about by summation (in DI_sum_) which absorbs all health deficits including reversible ones. Whole-of-life DIs were not significantly associated with elevated LV filling pressures, but the degree of health deficit accumulation between DI_60_64_ and any of the preceding time periods (DI_4–1_, DI_4–2_ and DI_4–3_) were. In a similar way, these step changes in health deficit accumulation but also those occurring earlier in life (DI_2-1_, DI_3-1_) were associated with lower LV systolic function at 60-64. These data suggest that regardless of one’s baseline health deficit burden, any rapid accumulation of multiple health deficits increases one’s risk of cardiac dysfunction in the senior years. It reinforces the point that even apparently mild *de novo* clinical issues should be taken seriously, to prevent them from having a long-term destabilizing effect on the myocardium. This is especially important given that with modern medicine, people are living longer, and previously deadly diseases are now being turned into chronic manageable problems contributing to multi-morbidity. It is already well-established that multi-morbidity erodes our well-being and quality of life, predisposing to further health deficit accumulation, premature death, mental health problems, negative health behaviours and reduced self-care conduct^15^. What the current work adds to the multi-morbidity knowledgebase, is this notion that rapid multiple health deficit accumulation is harmful for the heart.

A strength of the study is the implicit age homogeneity of birth cohort participants enabling age-matching at all subsequent assessments for DIs. Participants were exposed to similar risk factors and had access to similar diagnostic and treatment facilities over time. However, such secular effects may also have led to the underestimation of early-life DIs through underdiagnoses. Conversely, the burden of certain medical conditions may have been over-represented in early-life DIs compared to a modern-day population because treatments exist now that may not have been available then.

A limitation inherent to quantitative health deficit approaches^3^ is the equal weighting assigned to heterogenous health deficits that are unlikely to be straining the organism in a commensurate way. However, we performed a PCA and showed that the associations persisted for both LVmass_i_, and MCF_i_, even when the deficit index was formulated as a weighted average of health deficits. The associations were lost for EF and E/e’ because the weighted index diluted the contributions of cardiovascular deficits upon which EF and E/E’ strongly rely, as previously revealed by the sensitivity analysis (**Supplementary Table S5**). In addition, we did not account for the possibility of some health deficits spilling over (through causation or modulation) into other conditions. Another limitation is the potential of random zeros due to missing data, but we have shown that the prevalence of this was negligibly low in the analysed sample. The timing of echocardiographic assessment in the 7^th^ decade of life excludes participants that already passed away who might have had the highest DIs and most adverse cardiac phenotype. Moreover, selective follow-up could also be a potential confounder as changes in LV morphology could be a protective mechanism against mortality. There are recognized precision limitations in the GE Vivid I system used to capture all echocardiographic data in this study. Whilst the current data indicate a significant association between health deficit burden and echocardiographic parameters at 60-64 years, the evidence is insufficient to claim a causal line. Despite the association between high E/e’ and elevated LV filling pressure, there are instances where it is unreliable^16^. Although the prediction of normal and abnormal LV filling pressure is most reliable when the E/e’ is <8 or >15, cut-off of >13 was chosen to avoid sensitivity loss^17^.

## CONCLUSION

The accumulation of health deficits during the life-course associates with a maladaptive cardiac phenotype in older age characterized by hypertrophy and poorer function. It could be that health deficits accumulated during childhood, young adulthood, middle age and older age, insidiously strain the myocardium potentially paving the way to future cardiac dysfunction in susceptible individuals.

## Data Availability

NSHD data is available from: https://www.nshd.mrc.ac.uk/data.

## ACKNOWLEDGEMENTS

The authors would like to thank all the NSHD members for their participation and continuous engagement with follow-up and all NSHD scientific and data collection teams. The authors are also grateful to Andrew Wong and Adam Moore at the MRC Unit for Lifelong Health and Ageing at UCL for their assistance with NSHD data access.

